# Association between work attendance when experiencing fever or cold symptoms and company characteristics and socioeconomic status in the COVID-19 pandemic in Japanese workers: a cross-sectional study

**DOI:** 10.1101/2021.09.13.21263476

**Authors:** Kazuyoshi Mizuki, Makoto Okawara, Ayako Hino, Hajime Ando, Tomohisa Nagata, Seiichiro Tateishi, Mayumi Tsuji, Shinya Matsuda, Yoshihisa Fujino, for the CORoNaWork project

## Abstract

**Objective:** This study investigated the association between attending work while experiencing fever or cold symptoms and workers’ socioeconomic background and company characteristics during the COVID-19 pandemic.

**Methods:** A cross-sectional online survey was performed. Of a total of 33,302 participants, 3,676 workers who experienced fever or cold symptoms after April 2020 were included. The odds ratios (ORs) of attending work while sick associated with workers’ socioeconomic background and company characteristics were evaluated using a multilevel logistic model.

**Results:** The OR of attending work while sick associated with a lack of policy prohibiting workers from working when ill was 2.75 (95%CI: 2.28–3.20, P<0.001).

**Conclusion:** This study suggests that clear company policies on work and illness can be effective for preventing employees from attending work while sick.

## Introduction

Coronavirus disease 2019 (COVID-19), the disease caused by SARS-CoV-2, was first reported in China in December 2019. Since then, the disease has spread rapidly all over the world. The World Health Organization (WHO) officially declared COVID-19 a pandemic in March 2020,^1^ and the government of Japan first declared a state of emergency to minimize the rapid spread of COVID-19 in April 2020. The Japanese population was requested to wash their hands, wear masks, and avoid the three Cs: closed spaces, crowded places, and close-contact settings.

In addition to these countermeasures against infection in public places, the Japanese government emphasized the importance of implementing infection control measures in companies to prevent the spread of infection in society.^2,3^ This is because workers engage in numerous activities that can lead to spread of infection, including spending long hours in a small enclosed space with colleagues, engaging with customers, sharing business supplies, and talking with various people. Thus, the government and municipalities in Japan have required companies to take various infection control measures. Some of the most common COVID-19 measures Japanese companies have implemented include requesting that employees wear masks (94.2%), disinfect their hands (88.3%) and refrain from attending work when sick (84.0%), as well as disinfecting their workplaces (65.9%) and promoting telework (52.7%). ^4^ Government bodies have also emphasized the importance of measuring body temperature, which has been implemented by 72.8% of companies.

Countermeasures preventing employees with fever or cold symptoms from attending work are particularly important infection control measures in the workplace. Previous studies have reported that the most frequent symptom of COVID-19 is fever and that the virus is most infectious when symptoms are present.^5,6^ Thus, individuals who attend work while symptomatic for COVID-19 can easily infect their coworkers. For this reason, many companies have developed and informed their employees of policies related to working while symptomatic. In particular, some companies now require workers to check and report their body temperature before and at work, and have placed sensors at entrances to screen for people with fever.

Although most companies have introduced a policy prohibiting employees from attending work while sick, some employees continue to attend work regardless. There is little research on how often and why employees continue to attend work despite experiencing fever or cold symptoms during COVID-19 outbreaks. The phenomenon of continuing to work while sick is known as sickness presenteeism.^7^ Some possible reasons for sickness presenteeism are unstable employment, lack of paid leave, and a corporate tendency to praise attendance at work.^8^ In addition to sickness presenteeism, employees who continue to attend work while sick during the COVID-19 pandemic may have a low social status or poor work environment.

The objective of this study was to investigate the association between attending work while experiencing fever or cold symptoms and workers’ socioeconomic background and company characteristics during the COVID-19 pandemic in Japan.

## Methods

### Study design

We conducted a cross-sectional study on the health of workers in Japan during the COVID-19 epidemic using data from a prospective cohort study, the Collaborative Online Research on Novel coronavirus and Work study (CORoNaWork study).^9^ This study was conducted in accordance with the guidelines of the Declaration of Helsinki and was approved by the Ethics Committee of the University of Occupational and Environmental Health, Japan (reference No. R2-079 and R3-006). In addition, informed consent was obtained via a form on the internet.

A survey was performed from December 22 to 26, 2020, and included 33,302 workers between the ages of 20 and 65 years at the time of the study. Based on COVID-19 incidence data, participants were stratified by age, sex, occupation, and region. After excluding 6,051 who provided invalid responses, 27,036 participants were finally included. Details of the inclusion and exclusion criteria are provided in the protocol.^9^ Briefly, participants who completed the survey in extremely short response times, who were shorter than 140 cm, who weighed less than 30 kg, and who gave inconsistent answers to multiple identical questions were excluded.

### Assessment of attendance at work while sick

We asked the participants whether they attended work while experiencing fever or cold symptoms using a single question: “Did you attend work while experiencing fever or cold symptoms after April 2020?” Participants chose from one of the following options: had no symptoms, remained absent from work, worked at home, attended work with or without consulting the company. Participants who indicated they remained absent from work or worked at home were categorized as being absent while sick. Those who indicated they attended work with or without consulting the company were categorized as attending work while sick.

### Assessment of covariates

To examine the socioeconomic characteristics of individuals who attended work while sick, we inquired about the following items: job type (mainly desk work; mainly interpersonal communication; mainly physical work), company size (total number of workers in the company where the respondent mostly worked [self-employed business owner answered “1”]), marital status (married; unmarried; divorced/bereaved), annual equivalent household income (household income divided by the square root of household size), education (junior high school; high school; vocational school/college, university or graduate school), and degree of economic difficulty (very difficult; slightly difficult; comfortable).

To examine company characteristics, we inquired about the following items: whether or not the company had a policy requesting that employees refrain from attending work when they experience fever or cold symptoms, whether or not the worker received support from their supervisor or colleagues, and whether or not the worker had requested support from their company to continue working with a health condition. We used the Japanese version of the Job Content Questionnaire (JCQ) to assess the degree of support workers received from superiors and colleagues.^10^ Each JCQ item is scored on a four-point scale from 1 (*strongly disagree*) to 4 (*strongly agree*) on the presence of support. Support from a supervisor or colleagues was assessed from a total of four items, with total scores ranging from 4 to 16. Each sub-scale of support was categorized into quartiles based on the distribution of scores in the sample.

As a community-level variable, we used the cumulative incidence of COVID-19 in the prefecture of residence from the time of the survey until one month before the survey. We obtained this information from the websites of public institutions.

### Statistical analyses

We estimated odds ratios (ORs) of attending work while ill using a multilevel logistic model nested by prefecture of residence. In the multivariate model, we adjusted for sex, age, job type, number of employees in the workplace, and company policy on attending work while ill. Stata (Stata Statistical Software: Release 16. College Station, TX: StataCorp LLC.) was used for the analysis. A p-value less than 0.05 was considered statistically significant.

## Results

Table 1 shows participant characteristics by absence or attendance at work while experiencing fever or cold symptoms. Of the 27,036 participants, 3,676 experienced fever or cold symptoms during the survey period. Among these, 2,853 (77.6%) were absent and 823 (22.4%) attended work while sick.

**Table 1.** Basic characteristics of the study subjects

|  | Absent while sick<br>(n=2,853) |  | Attended work while sick<br>(n=823) |  |
| --- | --- | --- | --- | --- |
|  | n | % | n | % |
| Sex, male | 1,271 | 44.5 | 387 | 47.0 |
| Marital status, married | 1,560 | 54.7 | 427 | 51.9 |
| Job type |  |  |  |  |
| Mainly desk work | 1,513 | 53.0 | 387 | 47.0 |
| Mainly interpersonal communication | 728 | 25.5 | 193 | 23.5 |
| Mainly labor | 612 | 21.5 | 243 | 29.5 |
| Annual equivalent household income (JPY) |  |  |  |  |
| 500,000–2,650,000 | 929 | 32.6 | 293 | 35.6 |
| 2,650,000–4,500,000 | 843 | 29.5 | 255 | 31.0 |
| >4,500,000 | 1,081 | 37.9 | 275 | 33.4 |
| Self-rated health, very good/good | 1,107 | 38.8 | 244 | 29.6 |
| Psychological distress (K6 ≥ 5) | 1,574 | 55.2 | 512 | 62.2 |
| Felt lonely (usually or always) | 405 | 14.2 | 170 | 20.6 |
| Requested support from company to continue working |  |  |  |  |
| No | 1,766 | 61.9 | 493 | 59.9 |
| Yes, but I have not received support | 714 | 25.0 | 254 | 30.9 |
| Yes, and I have received support | 373 | 13.1 | 76 | 9.2 |
| Degree of economic difficulty |  |  |  |  |
| Very difficult | 363 | 12.7 | 153 | 18.6 |
| Slightly difficult | 908 | 31.8 | 242 | 29.4 |
| Comfortable | 1,582 | 55.5 | 428 | 52.0 |
| Support from supervisor* | 11 (8-12)** |  | 10 (8-12)** |  |
| Support from colleagues* | 11 (9-12)** |  | 10 (8-12)** |  |

\* Support from supervisor or colleagues was evaluated using the Japanese version of the Japan Content Questionnaire (four items, total score range: 4-16). High scores indicate that the respondent strongly agreed that they received support.
\*\* Median and interquartile range.

Table 2 shows the ORs of attending work while sick associated with socioeconomic status estimated using a logistic model. In the multivariate analysis, having a job that mainly involves labor, working at a small to medium-sized company (10–99 employees or 100–999 employees), and being in a ‘very difficult’ financial situation were associated with attending work while ill. The adjusted OR (aOR) of attending work while ill among subjects whose job type involved mainly labor was 1.44 (95%CI: 1.19–1.74, p<0.001). Similarly, the aOR of attending work while ill among subjects who worked at small companies (10–99 employees) was 1.44 (95%CI: 1.10–1.87, p=0.007), and 1.46 (95%CI: 1.11–1.91, p=0.006) among those who worked at medium companies (100–999 employees). Finally, the aOR of attending work while ill among subjects with financial difficulties was 1.29 (95%CI: 1.03–1.62, p=0.027).

**Table 2.** Odds ratio of attending work while experiencing fever or cold symptoms associated with socioeconomic status

|  | Age-sex-adjusted |  |  |  | Multivariate* |  |  |  |
| --- | --- | --- | --- | --- | --- | --- | --- | --- |
|  | OR | 95% CI |  | p | aOR | 95% CI |  | p |
| Job type |  |  |  |  |  |  |  |  |
| Mainly desk work | reference |  |  |  | reference |  |  |  |
| Mainly interpersonal communication | 1.05 | 0.86 | 1.27 | 0.655 | 1.01 | 0.83 | 1.23 | 0.923 |
| Mainly labor | 1.55 | 1.29 | 1.87 | <0.001 | 1.44 | 1.19 | 1.74 | <0.001 |
| Company size |  |  |  |  |  |  |  |  |
| 2-9 | reference |  |  |  | reference |  |  |  |
| 10-99 | 1.22 | 0.95 | 1.58 | 0.125 | 1.44 | 1.10 | 1.87 | 0.007 |
| 100-999 | 1.09 | 0.84 | 1.40 | 0.529 | 1.46 | 1.11 | 1.91 | 0.006 |
| ≥1,000 | 0.84 | 0.65 | 1.09 | 0.193 | 1.22 | 0.92 | 1.61 | 0.162 |
| Marital status |  |  |  |  |  |  |  |  |
| Married | reference |  |  |  | reference |  |  |  |
| Unmarried | 1.30 | 0.99 | 1.70 | 0.058 | 1.20 | 0.91 | 1.58 | 0.207 |
| Divorced/widowed | 1.12 | 0.95 | 1.34 | 0.186 | 1.08 | 0.90 | 1.28 | 0.421 |
| Annual equivalent household income (JPY) |  |  |  |  |  |  |  |  |
| 500,000-2,650,000 | 1.26 | 1.04 | 1.52 | 0.016 | 1.06 | 0.87 | 1.29 | 0.586 |
| 2,650,000-4,500,000 | 1.19 | 0.98 | 1.44 | 0.078 | 1.09 | 0.89 | 1.33 | 0.401 |
| >4,500,000 | reference |  |  |  | reference |  |  |  |
| Education |  |  |  |  |  |  |  |  |
| Junior high school | 1.71 | 0.92 | 3.18 | 0.091 | 1.19 | 0.63 | 2.27 | 0.594 |
| High school | 1.23 | 1.03 | 1.47 | 0.024 | 1.13 | 0.93 | 1.36 | 0.215 |
| Vocational school/college, university, graduate school | reference |  |  |  | reference |  |  |  |
| Degree of economic difficulty |  |  |  |  |  |  |  |  |
| Very difficult | 1.55 | 1.25 | 1.93 | <0.001 | 1.29 | 1.03 | 1.62 | 0.027 |
| Slightly difficult | 0.98 | 0.82 | 1.17 | 0.857 | 0.91 | 0.76 | 1.10 | 0.335 |
| Comfortable | reference |  |  |  | reference |  |  |  |

\* The multivariate model was adjusted for age, sex, job type, company size, company policy on attending work while ill

Table 3 shows the ORs of attending work while sick associated with the company environment estimated using a logistic model. In the age-sex-adjusted model, a lack of a policy requesting that employees refrain from attending work while ill was significantly associated with attending work while ill (OR=2.70, 95%CI: 2.28–3.20, p<0.001). A signification association was also observed in the multivariate analysis (aOR=2.75, 95%CI: 2.30–3.28, p<0.001). Lack of support from superiors or colleagues was likewise significantly associated with attending work while ill in the multivariate analysis. The aOR of attending work while ill among subjects with lower levels of support from supervisors (JCQ: 4–7) was 1.70 (95%CI: 1.36–2.12, p<0.001). Meanwhile, the aOR of attending work while ill among subjects with lower levels of support from colleagues (JCQ: 4–8) was 1.47 (95%CI: 1.19–1.80, p<0.001).

**Table 3.** Odds ratio of attending work while experiencing fever or cold symptoms associated with company characteristics

|  | Age-sex-adjusted |  |  |  | Multivariate* |  |  |  |
| --- | --- | --- | --- | --- | --- | --- | --- | --- |
|  | OR | 95%CI |  | p | aOR | 95%CI |  | p |
| Policy requesting that employees refrain from attending work while ill |  |  |  |  |  |  |  |  |
| Yes | reference |  |  |  | reference |  |  |  |
| No | 2.70 | 2.28 | 3.20 | <0.001 | 2.75 | 2.30 | 3.28 | <0.001 |
| Support from supervisor** |  |  |  |  |  |  |  |  |
| Very Low (4–7) | 2.11 | 1.70 | 2.61 | <0.001 | 1.70 | 1.36 | 2.12 | <0.001 |
| Low (8–10) | 1.34 | 1.10 | 1.62 | 0.003 | 1.15 | 0.95 | 1.40 | 0.157 |
| High (11) | 1.41 | 1.08 | 1.84 | 0.012 | 1.35 | 1.03 | 1.78 | 0.030 |
| Very High (12–16) | reference |  |  |  | reference |  |  |  |
| Support from colleagues** |  |  |  |  |  |  |  |  |
| Very Low (4–8) | 1.74 | 1.43 | 2.13 | <0.001 | 1.47 | 1.19 | 1.80 | <0.001 |
| Low (9–10) | 1.28 | 1.04 | 1.59 | 0.022 | 1.16 | 0.93 | 1.44 | 0.194 |
| High (11) | 1.28 | 1.01 | 1.62 | 0.042 | 1.26 | 0.99 | 1.60 | 0.058 |
| Very High (12–16) | reference |  |  |  | reference |  |  |  |
| Requested support from company to continue working |  |  |  |  |  |  |  |  |
| No | reference |  |  |  | reference |  |  |  |
| Yes, but I have not received support | 1.27 | 1.07 | 1.51 | 0.008 | 1.06 | 0.88 | 1.27 | 0.558 |
| Yes, and I have received support | 0.73 | 0.56 | 0.95 | 0.021 | 0.72 | 0.55 | 0.94 | 0.016 |

\* The multivariate model was adjusted for age, sex, job type, company size, company policy on attending work while ill
\*\* Support from supervisor or colleagues was evaluated using the Japanese version of the Japan Content Questionnaire (four items, total score range: 4–16). High scores indicate that the respondent strongly agrees that they received support.

## Discussion

This study showed that about 20% of workers in Japan who experienced fever or cold symptoms attended work during the COVID-19 pandemic. We found that a lack of a company policy requesting that employees refrain from attending work when sick increased inappropriate attendance. In addition, vulnerable socioeconomic status and support from colleagues or supervisors were also associated with attending work when sick.

Even during an infectious disease pandemic, some individuals continue to engage in social activities despite showing disease-related symptoms. A study during the 2009 swine-origin influenza A (H1N1) pandemic reported that 36% of diagnosed cases attended work with a fever.^11^ In our present study, 22% of Japanese workers who experienced fever or common cold symptoms reported attending work while symptomatic. An earlier study during the COVID-19 pandemic reported that 32.3% of workers in London^12^ and 62.2% in Japan^13^ attended work even if they showed symptoms. Such behaviors can spread infection because workplaces are often crowded and interactions between workers provide ample opportunity for infection. It is thus necessary to implement countermeasures to prevent individuals from attending work when they are sick.

This study showed that a lack of a policy prohibiting workers from attending work while sick increased the number of individuals who did so. Because workplaces inherently operate through a chain of command, a clear policy can be beneficial for reducing inappropriate attendance at work. To effectively enforce such policies, many companies additionally provide their employees with access to time off while ill. During the influenza A (H1N1) epidemic, a strategy by companies to provide their employees paid time off while symptomatic reduced inappropriate attendance.^14^ Even after adjusting for job type and company size, the present study showed that introducing a policy requesting that workers refrain from attending work when symptomatic also reduced employee attendance while sick. This result suggests that company policies are effective regardless of worker job types or company size. While many companies have introduced a policy requesting that individuals refrain from attending work while sick during the COVID-19 pandemic, ^4^ small and medium-sized enterprises (SMEs) are less likely to introduce such a policy compared to large companies.^15^ SMEs are unable to establish such a policy because they experience greater difficulty guaranteeing employees an income and employment.^16^ Our results provide evidence that all companies, including SMEs, should implement more infection control measures, including requests for workers to stay home while ill.

In this study, we found that worker attendance at work while sick increased among those with lower levels of support from their supervisors and coworkers. This finding is in line with previous studies on sickness presenteeism. Previous studies have shown that a supportive work environment, including support from both supervisors and coworkers, can reduce the incidence of sickness presenteeism because employees are less likely to regard absenteeism due to sickness as inappropriate.^17,18^ A similar trend has been observed in research conducted during the COVID-19 pandemic, which found that support from supervisors or colleagues decreased the prevalence of sickness presenteeism.^19^ The COVID-19 pandemic has reduced communication among workers and increased loneliness, making support from supervisors and coworkers even more critical during this period.^20^

This study also showed that workers tended to attend work while ill if they had unstable socioeconomic status. A previous study showed that those with unstable socioeconomic status reported an intention to work despite COVID-19 infection, including those with low income, lack of insurance, and lack of food security.^21^ Personal factors contributing to sickness presenteeism include the inability to say no to work and financial difficulties, while work factors include excessive workload and lack of replacements.^8,22,23^ The importance of providing support for workers in vulnerable socioeconomic situations is increasingly emphasized, backed by the results of the present study conducted during the COVID-19 pandemic. Thus, to prevent workers with cold symptoms from attending work in the future, financial support and rights to paid leave are imperative, especially for those in an unstable socioeconomic situation.

The present study indicates that companies should establish a policy that prevents employees from attending work when they are sick. Inappropriate attendance can facilitate the spread infection in society through the occurrence of clusters in the workplace and contact with an unspecified number of people while commuting. Nevertheless, some people continue to attend work while ill, especially those in unstable socioeconomic situations and those working in SMEs. The present study showed that a policy that prevents attendance at work while an employee is ill is effective even after adjusting for individual socioeconomic conditions and the size of the workplace, indicating that such a policy can be useful regardless of an individual’s socioeconomic status or company size. Financial support from the government would further aid the implementation of such a policy in small-scale establishments.

This study has several limitations. First, because this was an Internet-based study, and it is uncertain whether the results are generalizable. However, we selected participants by occupation, region, and prefecture based on infection incidence to minimize bias among the participants. Second, company policies on COVID-19 identified in this study were based on participant reports; it is possible that some participants were unaware of their company’s policies.

In conclusion, we showed that the absence of a company policy requesting that employees refrain from attending work while sick is significantly associated with increased attendance while experiencing fever or cold symptoms. We also showed that individuals with an unstable socioeconomic status and lower level of support from their supervisors and coworkers are more likely to attend work while sick. In the future, it is crucial to encourage more companies to establish policies that request employees to refrain from attending work while sick and to support those with unstable socioeconomic status.

## Data Availability

Data not available due to ethical restrictions

## Acknowledgments

The current members of the CORoNaWork Project, in alphabetical order, are as follows: Dr. Yoshihisa Fujino (present chairperson of the study group), Dr. Akira Ogami, Dr. Arisa Harada, Dr. Ayako Hino, Dr. Hajime Ando, Dr. Hisashi Eguchi, Dr. Kazunori Ikegami, Dr. Kei Tokutsu, Dr. Keiji Muramatsu, Dr. Koji Mori, Dr. Kosuke Mafune, Dr. Kyoko Kitagawa, Dr. Masako Nagata, Dr. Mayumi Tsuji, Ms. Ning Liu, Dr. Rie Tanaka, Dr. Ryutaro Matsugaki, Dr. Seiichiro Tateishi, Dr. Shinya Matsuda, Dr. Tomohiro Ishimaru, and Dr. Tomohisa Nagata. All members are affiliated with the University of Occupational and Environmental Health, Japan.

## Clinical significance

This study showed that a lack of a policy requesting that employees refrain from attending work while ill was associated with attendance at work while experiencing fever or cold symptoms. Thus, such a policy is a beneficial countermeasure for companies to prevent the spread of infection.

